# Change of dominant strain during dual SARS-CoV-2 infection

**DOI:** 10.1101/2020.11.29.20238402

**Authors:** AE Samoilov, VV Kaptelova, AY Bukharina, OY Shipulina, EV Korneenko, AV Lukyanov, AA Grishaeva, AA Ploskireva, AS Speranskaya, VG Akimkin

**Affiliations:** Central Research Institute of Epidemiology of the Federal Service on Customers’ Rights Protection and Human Well-being Surveillance, Moscow, Russia

## Abstract

**Background:** The effect of SARS-CoV-2 mutations and viral load on the severity of COVID-19 is not well understood. The possibility of reinfection with SARS-CoV-2 has already been reported, but dual infection with SARS-CoV-2 is poorly described and is currently under discussion. We describe a study of two strains of SARS-CoV-2 detected in the same patient during the same disease presentation.

**Methods:** Two nasopharyngeal swabs were obtained eight days apart from the patient in their 90s, diagnosed with lobar pneumonia (J18.1). Both tests were positive for SARS-CoV-2 with high viral load (Ct = 13). We have performed high-throughput sequencing of SARS-CoV-2 genomes from both swabs.

**Findings:** Genomic analysis of SARS-CoV-2 revealed the presence of two genetically distant strains in both swabs. Detected strains belong to different phylogenetic clades (GH and GR) and differ in the seven nucleotide positions. The relative abundance of strains was 70% (GH) and 30% (GR) in the first swab, and 3% (GH) and 97% (GR).

**Interpretation:** Our findings suggest that the patient was infected by two genetically distinct SARS-CoV-2 strains at the same time. One of the possible explanations is that the second infection occurred in the hospital. Change of the dominant strain ratio during disease manifestation could be explained by the advantage or higher virulence of the strain belonging to the clade GR.

## Introduction

Dual infection is a phenomenon where an individual is simultaneously infected with two or more strains of the same virus. It can affect host immune responses and result in increased fitness of the viral population. A number of cases when individuals were infected with more than one strain of HIV were identified in the last years [1-3]. The findings of dual infections were reported for influenza viruses [4], the Epstein-Barr virus [5], and other viruses. Cases of SARS-CoV-2 reinfection are described in the scientific literature [6 - 8]; however, there are almost no reports of double infection with SARS-CoV-2 with the exceptions of in two works from Iraq [9] and Switzerland [10]. Here, we present a case report of an individual with two genetically distinct SARS-CoV-2 strains during the same illness manifestation. These strains were classified into different phylogenetic clades: GH and GR. We demonstrated that the abundance of the strains could change significantly over time.

## Materials and methods

A patient in their 90s with a medical history of chronic persistent atrial fibrillation, heart failure, and hypertension was hospitalized with fever (38 °C) with the admission diagnosis of lobar pneumonia (J18.1). Oropharyngeal swab obtained on the next day (sample 1) tested positive for SARS-CoV-2 (cycle threshold, Ct = 13, measured using AmpliSens® Cov-Bat-FL assay kit). Five days later, the patient was transferred to the ICU of the hospital, specialized in the treatment of COVID-19 patients. During the observation period, the patient’s condition progressively worsened. Oxygen saturation ranged from 70% without oxygen support to 92 - 98% with oxygen support with an oxygen mask. Repeated oropharyngeal swab for SARS-CoV-2 (sample 2), which was taken eight days after the first swab, also tested positive with high viral load (Ct = 13, measured using AmpliSens® Cov-Bat-FL assay kit). After five days of ICU treatment, the patient died. The cause of death was a coronavirus infection, which progressed unfavorably due to premorbid status.

RNA was extracted using the QIAamp Viral RNA Mini Kit (QIAGEN, Germany) according to the manufacturer’s instructions. Reverse transcription reaction was performed using 10 μL of the RNA samples, random hexanucleotide primers, and Reverta-L kit (AmpliSens, Russia) according to the manufacturer’s instructions. Libraries for whole-genome sequencing of SARS-CoV-2 were prepared using single-strand cDNA, and SCV-2000bp protocol described previously [11]. In brief, we amplified four pools of ∼2000 bp-long fragments covering the whole SARS-CoV-2 genome. The obtained PCR products were purified, mixed equally, and sheared in microTUBE-50 AFA Fiber Screw-Cap (PN 520166) using Covaris M220 (Covaris, Woburn, MA). Libraries were constructed using Y-shaped adapters compatible with Nextera XT Index Kit and amplified with 8 cycles using oligos from Nextera XT Index Kit and Q5 High-Fidelity DNA Polymerase (New England BioLabs, NEB).

Libraries for total RNA sequencing were prepared using the NEBNext® Ultra™ II Directional RNA Second Strand Synthesis Module E7550 (according to the manufacturer’s instructions, New England BioLabs, NEB). Double-stranded cDNA was sheared in microTUBE-50 AFA Fiber Screw-Cap (PN 520166) using Covaris M220 (Covaris, Woburn, MA) using the following settings: peak incident power - 75W, duty factor - 10%, cycles per burst - 200, treatment Time - 40 s, temperature - 20 °C, sample volume - 50 µl. Libraries were constructed with NEBNext® Ultra™ II End Repair/dA-Tailing Module (E7546), NEBNext Ultra II Ligation Module (E7595) according to the manufacturer instructions (New England BioLabs, NEB). Amplification of libraries was performed with Q5 High-Fidelity DNA Polymerase (M0491) using NEBNext® Multiplex Oligos for Illumina® (Index Primers Set 2, E7500) in total 25 μL according to the manufacturer instructions (New England BioLabs, NEB) with 10 cycles of amplification.

Size selection of the libraries was performed using Agencourt AMPure XP (Beckman Coulter, Danvers, MA, USA). Quality and fragment length distribution of the obtained libraries were evaluated with Agilent Bioanalyzer 2100 (Agilent Technologies, USA). Sequencing was performed on Illumina HiSeq 1500 with HiSeq PE Rapid Cluster Kit v2 and HiSeq Rapid SBS Kit v2 (500 cycles).

Raw reads from amplicon libraries and total RNA libraries were processed as described in [11]. We performed adapter and quality trimming with Trimmomatic [12], removed PCR primers with cutadapt (for amplicon libraries) [13], and mapped reads to the reference sequence (strain hCoV-19/Wuhan/WIV04/2019, GISAID accession ID EPI_ISL_402124) using bowtie2 [14]. After that, we filtered out reads with low mapping quality using SAMtools [15], performed base-calling with GATK [16], filtered gvcf files were with BCFtools [17]. Finally, the consensus sequences were obtained with BEDTools [18]. The validity of the resulting sequence was verified manually by visual inspection of mapped reads. Areas with coverage lower than 5 were masked with NNN. The frequency of nucleotides over the SARS-CoV-2 genome was analyzed using the Rsamtools package. All of the SARS-CoV-2 strains were aligned using mafft [19], the phylogenetic tree was built using IQtree 2 [20] using the GTR model.

## Results

We have performed the sequencing of two swab samples obtained from the same patient eight days apart using the SCV-2000bp protocol and Illumina sequencing. The sequencing of sample 1 yielded 1.1 M paired-end raw reads. After quality filtration and PCR primer trimming, 837 thousand reads remained, 99.87% of which mapped to the reference SARS-CoV-2 genome strain hCoV-19/Wuhan/WIV04/2019 (GISAID accession ID EPI_ISL_402124). The mapping of trimmed reads to a reference sequence revealed seven heterogeneous positions (see Fig. 1 for an example).

**FIGURE 1.**
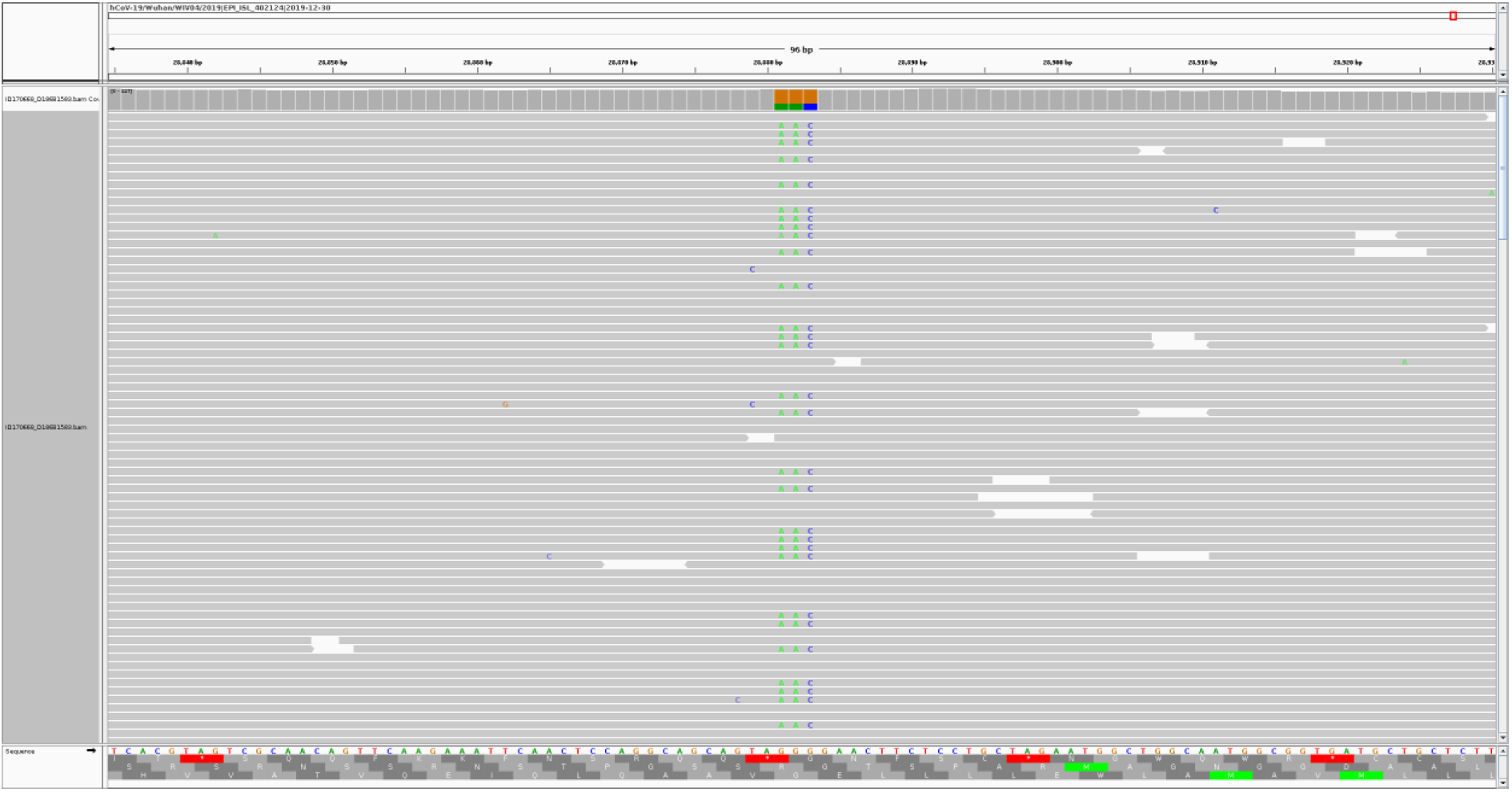
Heterogeneity in mapped Illumina reads in the first sample 1. 66 % of the reads match the reference SARS-CoV-2 sequence strain hCoV-19/Wuhan/WIV04/2019 (GISAID accession ID EPI_ISL_402124), 34% of reads have GGG->AAC substitution at positions 28881-28883.

The sequencing of sample 2 yielded 3.9 M paired-end raw reads. After quality filtration and PCR primer trimming, 3.7 M reads remained, 99.93% of which mapped to the reference SARS-CoV-2 genome strain hCoV-19/Wuhan/WIV04/2019 (GISAID accession ID EPI_ISL_402124). We analyzed the mapped reads and found the same heterogeneity at the same positions as in Sample 1, but at a much lower level.

We interpreted our observations as the simultaneous presence of two SARS-CoV-2 strains in the same patient’s samples. After obtaining consensus genomic sequence from the dominant strain from the less heterogeneous second sample (hereafter called strain 2), it became possible to unambiguously reconstruct genomic sequences of the strain prevalent in the first sample (hereafter called strain 1). Read coverage at the genomic positions differentiating strain 1 from strain 2 varied from 288 to 16000 in sample 1 and from 950 to 64000 in sample 2. The relative abundance of strains 1 and 2 in both time points was assessed by averaging the relative coverage of heterogeneous positions (Figure 2A) and amounted to roughly 69% and 31% in the first sample and 3% and 97% in the second sample, respectively (Fig. 2C). We found that strain 1 was dominant in sample 1, and strain 2 became dominant in sample 2.

**FIGURE 2.**
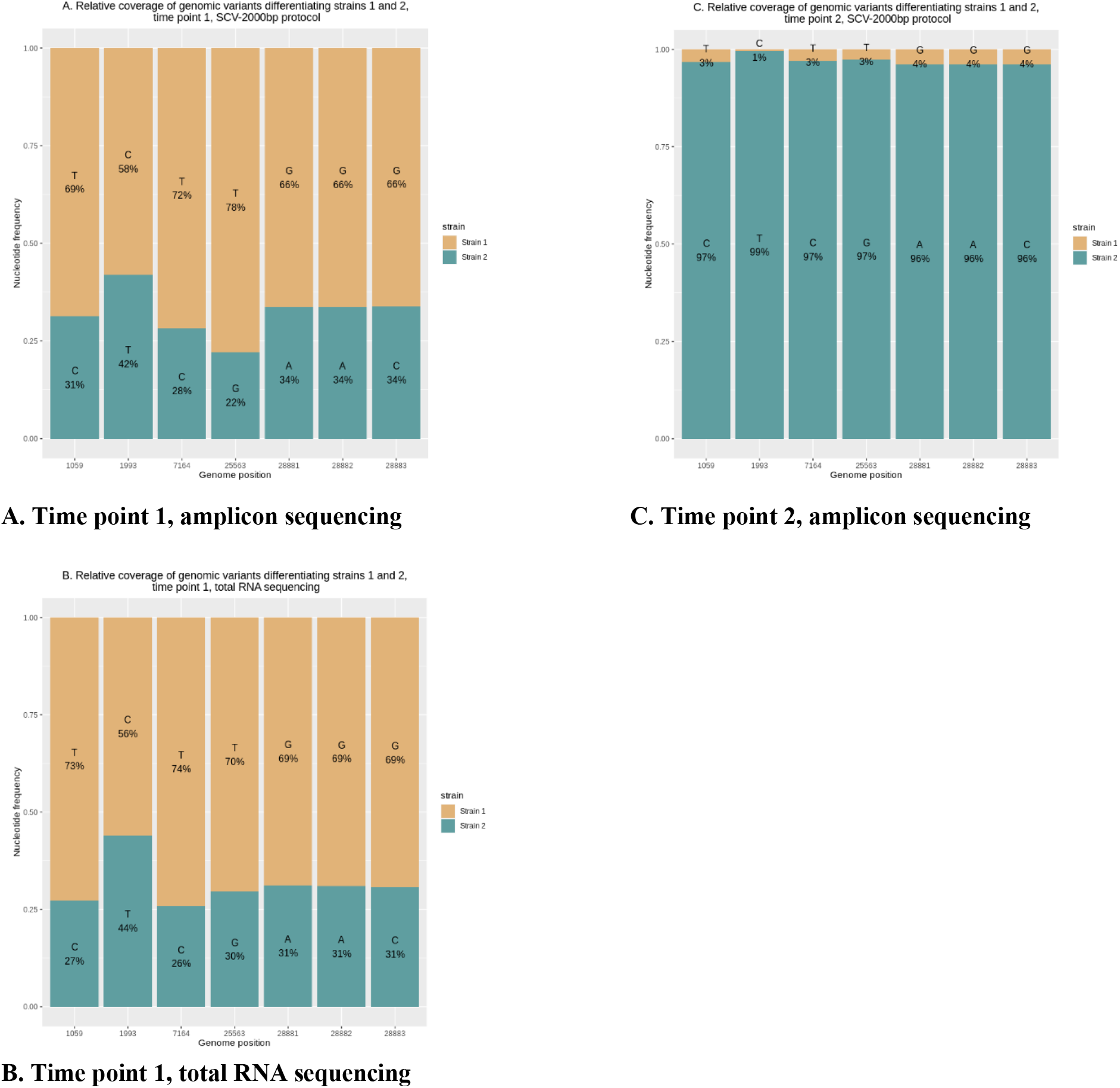
Relative coverage of SARS-CoV-2 genomic variants, which differentiate strains 1 and 2, at the time points 1 and 2. Genomic positions are at the X-axis, their relative frequencies are at Y-axis. Sample 1, sequencing using: (a) genome fragments amplification (SCV-2000bp protocol), (b) total RNA library. Sample 2 (collected eight days later), sequencing using: (c) genome fragments amplification (SCV-2000bp protocol).

The resulting sequences are available at GISAID with accession numbers EPI_ISL_610237 (hCoV-19/Russia/CRIE170668/2020, the dominant strain from Sample 1) and EPI_ISL_610238 (hCoV-19/Russia/CRIE300223/2020, the dominant strain from Sample 2, collected eight days after the first swab).

Heterogeneity in the sequence reads can also be explained by sequencing artifacts arising from the polymerase errors, chimeric fragments generation during nucleic acid amplification, and contamination during RNA extraction and library preparation. To exclude the possibility that the observed heterogeneity is a mistake, we have isolated RNA from the original swabs for the second time and performed sequencing of libraries prepared from total RNA without any enrichment for both samples.

RNA-seq of sample 1 yielded 12.6 M paired-end 250 bp long reads. After quality filtration and PCR primer trimming, 12.2 M reads remained, 2.29% of which were mapped to the reference SARS-CoV-2 genome strain hCoV-19/Wuhan/WIV04/2019 (GISAID accession ID EPI_ISL_402124). We compared RNA-seq reads with reads obtained using the SCV-2000bp protocol (Fig. 2B). Roughly the same frequencies of alternative nucleotides prove that observed heterogeneity was not a result of a sequencing artifact and that DNA originating from two different SARS-CoV-2 strains is present in sample 1. Read coverage at the genomic positions differentiating strain 1 from strain 2 varied from 2100 to 4090 in sample 1 after total RNA sequencing.

RNA-seq of Sample 2 yielded 15.0 M paired-end raw reads. After quality filtration and PCR primer trimming, 14.6 M reads remained, and only 0.04 % of them were mapped to the reference SARS-CoV-2 genome strain hCoV-19/Wuhan/WIV04/2019 (GISAID accession ID EPI_ISL_402124), which was not enough to confirm the presence of a minor fraction of reads (about 3%) representing strain 1.

Comparison of strain 1 genomic sequence to all of the GISAID SARS-CoV-2 database (as of 11.11.2020) revealed that this sequence is unique to GISAID, the closest genomes having at least two mismatches compared to strain 1 genome. Out of 571 closest sequences, only three originated from Russia (EPI_ISL_428905, EPI_ISL_428875, EPI_ISL_428871, all of them were collected in March), most of the other genomes originated from the USA (402), Iceland (28), and Canada (26) and were collected in March-early April.

Comparison of strain 2 genomic sequence to all of the GISAID SARS-CoV-2 database (as of 11.11.2020) revealed 1062 genomic sequences with 100% identity to strain 2, 78 out of which originated from different regions of Russia, 10 of them were collected in Moscow (collection dates of which vary from late March to early April), including eight genomes obtained in our lab, as described in [11]. The latest genomes with 100% identity to strain 2 were collected in Saint-Petersburg in the middle of September (EPI_ISL_602339 and EPI_ISL_602340). Other genomes with 100% identity to strain 2 originated mostly from England (333), Portugal (121), and the USA (86) and were collected mainly in March and April.

Comparison of strain 1 and strain 2 genomic sequences with the reference strain hCoV-19/Wuhan/WIV04/2019 (GISAID accession ID EPI_ISL_402124) revealed four nucleotide mutations present in both of them (C241T in the non-coding region; C3037T, a synonymous substitution in NSP3 protein; C14408T, resulting in P323L mutation in NSP12 protein; A23403G, resulting in D614G mutation in spike protein), as well as four mutations, present only in strain 1 (C1059T, resulting in T85I mutation in NSP2 protein; T1993C, a synonymous substitution in NSP2 protein; C7164T, resulting in T1482I mutation in NSP3 protein; G25563T, resulting in Q57H in NS3 protein) and three mutations, present only in strain 2 (G28881A and G28882A, resulting in R203K mutation in N protein; G28881C, resulting in G204R mutation in N protein) (Table 1).

**Table 1.**
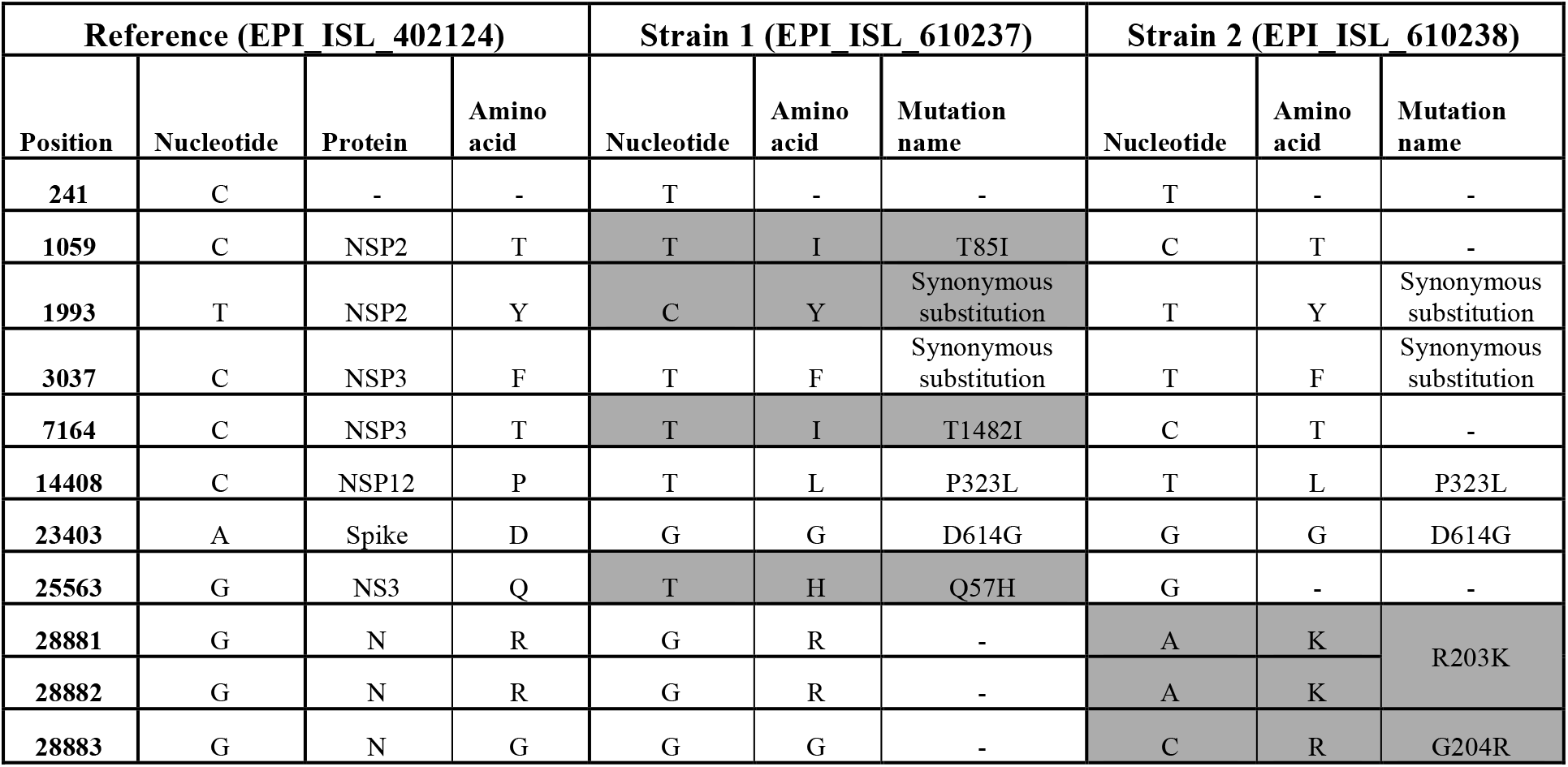
List of mutations present in strains 1 and 2. Mutations that differ strain 1 from strain 2 are marked by gray color.

Phylogenetic analysis was performed by building a tree of all of the available SARS-CoV-2 genomes from the samples collected in Russia. It has been revealed that strain 1 (EPI_ISL_610237) belongs to the GH clade, and strain 2 (EPI_ISL_610238) belongs to the GR clade (GISAID classification) (see Fig. 3).

**FIGURE 3.**
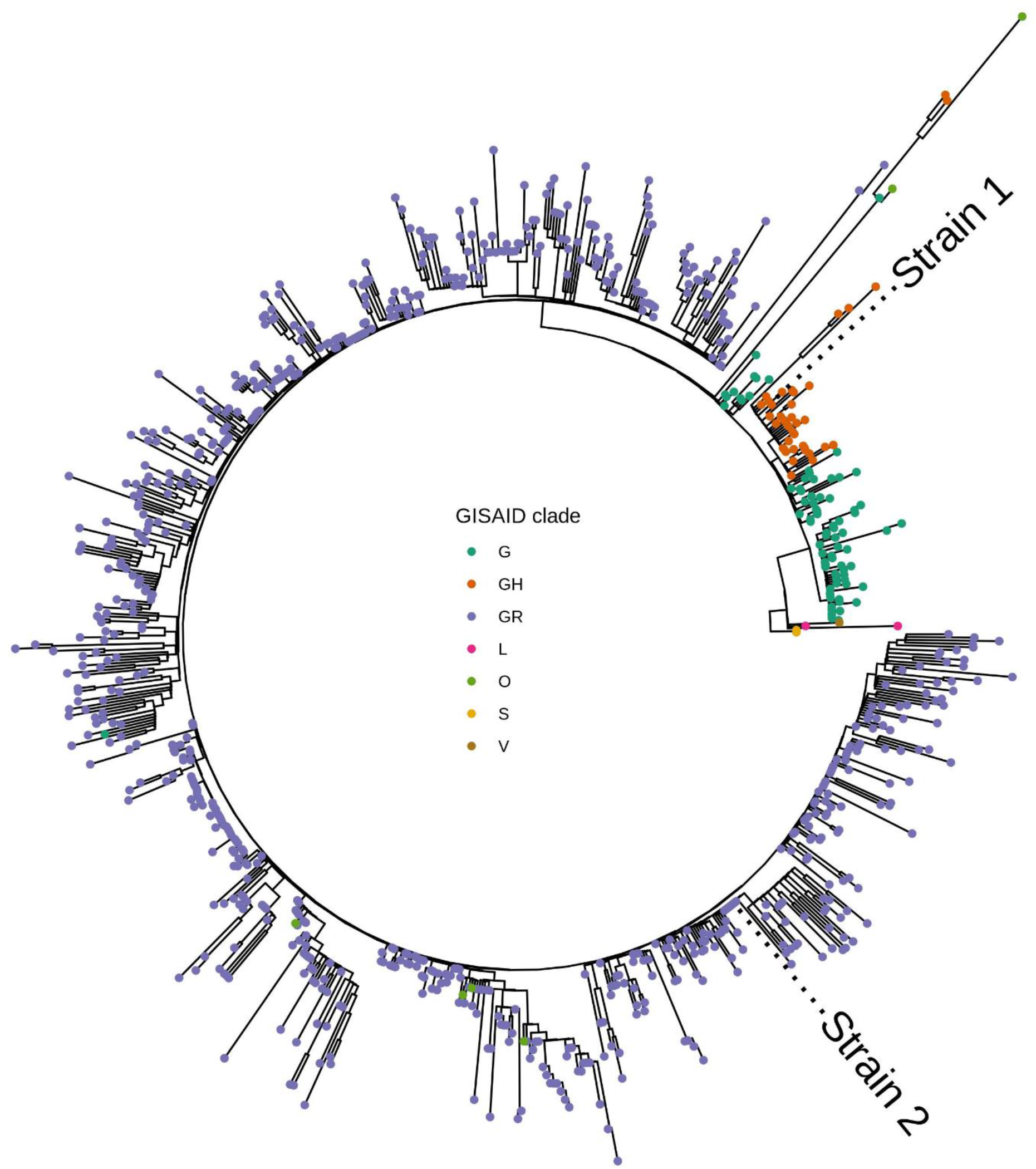
Phylogenetic tree of all of the available SARS-CoV-2 strains isolated in Russia (as of 11.11.2020). GISAID clade classification is represented by color. Tip labels mark positions of strain 1 (Russia/CRIE170668/2020, EPI_ISL_610237) and strain 2 (Russia/CRIE300223/2020, EPI_ISL_610238). Closely related to SARS-CoV-2 virus strain bat/Yunnan/RmYN02/2019 (EPI_ISL_412977) was used as a root (not shown).

## Discussion

Dual infection can affect host immune responses and result in increased fitness of the viral population. HIV dual infection contributes to rapid disease progression [1], increased viral load [2], and requires antiretroviral treatment effective against both viruses [3]. Meanwhile, despite a rapidly growing body of articles, there is almost no information about dual SARS-CoV-2 infection. To our knowledge, the possibility of SARS-CoV-2 double infection or within-patient genetic diversity was discussed in two works [9, 10]. Authors of both provided no information about patients’ medical history, viral subpopulation dynamics during disease progression, and almost no information about clade classification of SARS-CoV-2 analyzed strains.

Hashim et al. [9] utilized Sanger sequencing and found double peaks in short (795 bp) fragments of spike protein gene and interpreted it as the presence of double infection in all of the 19 analyzed samples. Unfortunately, the authors’ interpretation of Sanger sequencing data is questionable. Most of the electropherograms presented in the manuscript [9] undoubtedly consist of double peaks, but the authors also present too many electropherograms with low intensity of the minor peak. In conventional Sanger sequencing analysis, it is challenging to interpret the data when there is a low level of the second peak at the same nucleotide position. Minor variants can be reliably detected in the 20% mixture analyzed trace; however, in the analyzed traces from the 10% and 5% mixtures, the fragment is indistinguishable from the baseline noise [21].

The sensitivity of NGS is significantly superior to Sanger sequencing. Authors of the work [10] processed publicly available 3939 deeply sequenced SARS-CoV-2 genomes and 749 sequenced samples from Switzerland. The most interesting of their findings is that age was a statistically significant predictor of viral genetic diversity in the analyzed cohort. Authors have also discovered that Spike_D614G exhibits high intra-host diversity, with 29.7% of the publicly available cohort experiencing subclonal mutations with the different variants coexisting. In Switzerland data, the D614G variant is encoded by the second most diverse genomic position. The top 10 diverse positions from the publicly available data include eight positions from the ORF1ab gene and one position for genes S and ORF8. Top 10 diverse positions from Swiss data include four positions from ORF1ab gene, two positions from each of genes M and E, and one position from gene S.

The presence of two viral variants within the same patient might be associated with nosocomial infection. However, the patient spent only one day in the hospital until the collection of the first swab, in which we have detected two SARS-CoV-2 strains. The probability of getting a positive PCR test result in the early days after infection is rather small [22]. Therefore, our data can be interpreted either as an infection with two strains before admission to the hospital or as a rapid increase of hospital strain viral load to a detectable level due to the patient’s weakened immune system. The severity and rapid progression of the disease, along with unchangeably high viral load (Ct = 13), could be associated with either a change in the dominant strain of SARS-CoV-2 or the patient’s elderly age [23].

The mutation rate of SARS-CoV-2 inside the same host is a critical parameter for understanding viral evolution because it may become more infectious or more virulent. Choi et al. [24] described the case of persistent infection (over 150 days) accompanied by accelerated evolution of SARS-CoV-2 in an immunocompromised patient. On days 18 and 25, sequencing of SARS-CoV-2 genomes obtained from the patient revealed five amino acid substitutions compared to the reference, but later their number grew to over 20. The largest number of mutations was detected in the Spike protein, especially in the receptor-binding domain. We have not observed the appearance of new mutations, but strain 1, discussed in this work, is closely related to the strain obtained from the immunocompromised patient at day 18 (both of them possess mutations Spike_D614G, NS3_Q57H, NSP2_T85I, and NSP12_P323L, but differ in mutation NSP13_T115I present only in the strain obtained from the immunocompromised patient and NSP3_T1482I present only in the strain 1).

Our results show a drastic change in both strains’ abundance: the dominant strain from the first sample almost disappeared in the sample obtained over a week later. The strain dominating in the first sample belongs to GH clade, while the strain which prevailed in the second sample belongs to GR clade. Change of the dominant strain in the viral community can be explained by different relative fitness caused by advantageous or disadvantageous mutations that distinguish one strain from another. In our case, potentially disadvantageous mutations (present in strain 1, which decreased its abundance over time) include Q57H in NS3 protein, T85I in NSP2 protein, and T1482I in NSP3 protein. NS3_Q57H demarcates GH clade [25]. As of 11.11.2020, it occurs in 4.6 % (40 out of 874) of Russian SARS-CoV-2 sequences. Possible effects of this mutation were discussed in several papers. In a work by Gupta et al. [26], who used protein modeling, its effect was predicted as deleterious. Alam et al. discussed that it prevents ion permeability by constricting the channel pore more tightly, possibly reducing viral release and immune response [27]. In work by Wang et al. [28], the authors suggested that it can make the SARS-CoV-2 more infectious. The second mutation present only in strain 1, NSP2_T85I, is rare in Russia and occurs in 3.4% (30 out of 874) Russian SARS-CoV-2 genomes (as of 11.11.2020). This mutation also has predicted deleterious functional outcome [29]. Wang et al. [28] discussed that this mutation benefits from other mutations like Spike_D614G and NS3_Q57H and could strengthen infectivity. Finally, the mutation NSP3_T1482I is present in only 96 genomes submitted to GISAID (as of 11.11.2020), and it was never discussed in scientific literature to our knowledge.

Potentially advantageous mutations (present in the dominant strain in the second sample) include R203K and G204R in N protein. These mutations occur together as a result of the substitution of three consecutive nucleotides. The presence of these mutations demarcates clade GR [25]. Over 85% of Russian SARS-CoV-2 genomes submitted to GISAID (11.11.2020) belong to the GR clade and possess both of these mutations. According to different protein modeling approaches, these mutations either destabilize N protein [27] or have a neutral effect [26]. Several articles point out the association of this clade with higher mortality or significant prevalence among the group of severe disease or deceased patients and also higher prevalence in females and children compared to other clades [31].

Other mutations present in both strains are D614G in spike protein and P323L in NSP12. Spike_D614G is one of the most widely discussed mutations. Its presence defines G clade [25], it increases infectivity [32-34], mortality [35, 36], alters viral fitness [37], and, according to protein modeling, strengthens the folding stability of the spike protein [28]. It is present in 99.2 % of SARS-CoV-2 genomes obtained from Russia (as of 11.11.2020). NSP12_P323L is predicted to make the polymerase more rigid, which may increase the replication speed [28] and mutation rate [38, 39]. It is present in 97.9 % of Russian SARS-CoV-2 genomes (as of 11.11.2020).

In our work, we present a case of dual SARS-CoV-2 infection, which allowed us to compare the relative fitness of two genetically distant strains. The effect of SARS-CoV-2 dual infection on viral load and the severity or duration of COVID-19 is currently unknown. It is possible that presence of two different SARS-CoV-2 strains was a factor which lead to rapid progression of the patient’s disease and death. The importance of research aimed at cases similar to ours can hardly be overestimated because it allows insights into the molecular epidemiology of COVID-19 and can help detectpotentially advantageous mutations, which increase virulence and fitness of SARS-CoV-2.

## Conclusion

Our study shows the case of dual SARS-CoV-2 infection by two phylogenetically distant strains and the dynamic in the viral community. The strain that became dominant eight days after the first sample was collected belonged to the GR clade, which is most prevalent in Russia, which raises the question of whether strains from this clade have higher fitness compared to strains belonging to the other clades. We have also detected two potentially advantageous mutations (N_R203K and N_G204R, belonging to the strain which increased its abundance over time) and three potentially disadvantageous (NS3_Q57H, NSP2_T85I, and NSP3_T1482I, belonging to the strain which decreased its abundance over time).

## Data Availability

Data referred to in the manuscript is not available online.

## Acknowledgments

The authors would like to express their gratitude to the staff of the Molecular diagnostic laboratory of Federal Budget Institution of Science “Central Research Institute of Epidemiology” of The Federal Service on Customers’ Rights Protection and Human Well-being Surveillance for providing the anonymized samples of patients with COVID-19.

## Funding

This work was supported by the Ministry of Science and Higher Education of the Russian Federation within the framework of a grant in the form of a subsidy for the creation and development of the «World-class Genomic Research Center for Ensuring Biological Safety and Technological Independence under the Federal Scientific and Technical Program for the Development of Genetic Technologies», agreement No. 075-15-2019-1666.

## Notes

### Competing Interest Statement

The authors have declared no competing interest.

### Author Declarations

This research was approved by the local ethics committee of the Central Research Institute of Epidemiology Federal Service for Surveillance on Consumer Rights Protection and Human Wellbeing on 17.11.2020.

